# Modeling and Forecasting of COVID-19 New Cases in the Top 10 Infected African Countries Using Regression and Time Series Models

**DOI:** 10.1101/2020.09.23.20200113

**Authors:** Alemayehu Siffir Argawu

## Abstract

**Background:** COVID-19 total cases have reached 1,083,071 (83.5%) in the top 10 infected African countries (South Africa, Egypt, Morocco, Ethiopia, Nigeria, Algeria, Ghana, Kenya, Cameroon, and Côte d’Ivoire) from Feb 14 to Sep 6, 2020. Then, this study aimed to model and forecast of COVID-19 new cases in these top 10 infected African countries.

**Methods:** In this study, the COVID 19 new cases data have been modeled and forecasted using curve estimation regression and time series models for these top 10 infected African countries from Feb 14 to Sep 6, 2020.

**Results:** From July to August, the prevalence of COVID-19 cumulative cases was declined in South Africa, Cote d□Ivoire, Egypt, Ghana, Cameron, Nigeria, and Algeria by 31%, 26%, 22%, 20%, 14%, 12%, and 4%, respectively. But, it was highly raised in Ethiopia and Morocco by 41%, and 38% in this period, respectively. In Kenya, it was raised only by 1%. In this study, the cubic regression models for the ln(COVID-19 new cases) data were relatively the best fit for Egypt, Ethiopia, Kenya, Morocco, Nigeria and South Africa. And, the quadratic regression models for the data were the best fit for Cameroon, Cote d□Ivoire and Ghana. The Algeria data was followed the logarithmic regression model. In the time series analysis, the Algeria, Egypt, and South Africa COVID-19 new cases data have fitted the ARIMA (0,1,0), ARIMA (0,1,0), and ARIMA (0,1,14) models, respectively. The Cameroon, Cote d’Ivoire, Ghana, and Nigeria data have fitted the simple exponential smoothing models. The Ethiopia, Kenya, and Morocco data have followed the Damped trend, Holt, and Brown exponential smoothing models, respectively. In the analysis, the trends of COVID-19 new cases will be declined for Algeria and Ethiopia, and the trends will be constantan for Cameroon, Cote d’Ivoire, Ghana and Nigeria. But, it will be raised slightly for Egypt and Kenya, and significantly for Morocco and South Africa from September 7 to October 6, 2020.

**Conclusion:** This study was conducted with the current measures; the forecasts and trends nobtained may differ from the number of cases that occur in the future. Thus, the study finding should be useful in preparedness planning against further spread of the COVID-19 epidemic in African countries. And, the researcher recommended that as many countries continue to relax restrictions on movement and mass gatherings, and more are opening their airspaces, and the countries’ other public and private sectors are reopening. So, strong appropriate public health and social measures must be instituted on the grounds again.

## 1. Introduction

On 31 December 2019, the WHO China country office was informed of cases of pneumonia unknown etiology (unknown cause) detected in Wuhan city, Hubei province of China. On 3 January 2020, a total of 44 case-patients with pneumonia of unknown etiology were reported to WHO by the national authorities in China. On 7 January 2020, the Chinese authorities identified a new type of coronavirus. Then, it has quickly spread to become a global pandemic [1,2].

On 6 September 2020, after eight months, it was distributed to 215 countries and territories around the world. As of 6 September 2020 on 1:00 pm, 27,087,151 total cases of COVID-19 have been reported, including 884,110 total deaths with 3.3% of fatality rate and 19,208,336 total recovered with 70.9% recovered rate and also 6,994,705 active cases with 25.8% rate as Worldometers information provided globally. In Africa as this report showed that the total cases was 1,297,434 with 4.8% covered from the world, total deaths was 31,131 with 2.4% of fatality rate, total recovered was 1,033,859 with 79.7% of recovery rate, and active cases was 232,444 with 17.9% rate [3-5]. And, the current Worldometer information statistics were summarized clearly on table 1 below.

**Table 1:** Statistics on COVID-19 Cases of the countries between Feb 14 and Sep 6, 2020.

| World/Africa | Total Cases |  | Total Deaths |  | Total Recovered |  | Total Active Cases |  |
| --- | --- | --- | --- | --- | --- | --- | --- | --- |
|  | Counts | % | Counts | % | Counts | % | Counts | % |
| World | 27,087,151 | 100 | 884,110 | 3.3 | 19,208,336 | 70.9 | 6,994,705 | 25.8 |
| Africa | 1,297,434 | 4.8 | 31,131 | 2.4 | 1,033,859 | 79.7 | 232,444 | 17.9 |
| South Africa (1<sup>st</sup>) | 636,884 | 49.1 | 14,779 | 2.3 | 561,204 | 88.1 | 60,901 | 9.6 |
| Egypt (2<sup>nd</sup>) | 99,712 | 7.7 | 5,511 | 5.5 | 77,208 | 77.4 | 16,993 | 17.0 |
| Morocco (3<sup>rd</sup>) | 70,160 | 5.4 | 1,329 | 1.9 | 53,929 | 76.9 | 14,902 | 21.2 |
| Ethiopia (4<sup>th</sup>) | 57,466 | 4.4 | 897 | 1.6 | 20,776 | 36.2 | **35,793** | **62.3** |
| Nigeria (5<sup>th</sup>) | 54,905 | 4.2 | 1,054 | 1.9 | 42,922 | 78.2 | 10,929 | 19.9 |
| Algeria (6<sup>th</sup>) | 46,071 | 3.6 | 1,549 | 3.4 | 32,481 | 70.5 | 12,041 | 26.1 |
| Ghana (7<sup>th</sup>) | 44,777 | 3.5 | 283 | 0.6 | 43,693 | 97.6 | 801 | 1.8 |
| Kenya (8<sup>th</sup>) | 35,020 | 2.7 | 594 | 1.7 | 21,158 | 60.4 | 13,268 | 37.9 |
| Cameroon (9<sup>th</sup>) | 19,604 | 1.5 | 415 | 2.1 | 18,448 | 94.1 | 741 | 3.8 |
| Cote d'Ivoire (10<sup>th</sup>) | 18,472 | 1.4 | 119 | 0.6 | 17,323 | 93.8 | 1,030 | 5.6 |
| Total | 1,083,071 | 83.5 | 26,530 | 85.2 | 889,142 | 86.0 | 167,399 | 72.0 |

Source: https://www.worldometers.info/coronavirus/.

As of 1 September 2020, a cumulative total of 1,259, 817 COVID-19 cases were reported in the African region. In the region, there was a 3% total cases increment only in six days from September 1 to 6. And as of 6 September 2020, South Africa has registered more than 49% (636, 884), of all reported confirmed cases in the region. The other countries that have reported large numbers of cases were Egypt 7.7 % (99,712), Morocco 5.4% (70,160), Ethiopia 4.4% (57,466), Nigeria 4.2% (54,905), Algeria 3.6% (46,071), Ghana 3.5% (44,777), Kenya 2.7% (35, 020), Cameroon 1.5% (19,604) and Côte d’Ivoire 1.4% (18,472). These top 10 infected countries collectively have accounted 83.5% (1,083,071) of all reported cases in African region [3-5].

And as of 6 September 2020, the Worldometer updated information and the summarized table below have also shown that Egypt had recorded the highest fatality rate (5.5% with 5,511 deaths) in Africa. And the next 9 countries that have the highest fatality rates were Algeria (3.4% with 1,549 deaths), South Africa (2.3% with 14,779 deaths), Cameroon (2.1% with 415 deaths), Morocco and Nigeria have equal rates (1.9% with 1,329 and 1,054 deaths, respectively), Kenya 2 (1.7% with 594 deaths), Ethiopia (1.6% with 894 deaths), Ghana and Cote d□Ivoire have equal rates (0.6% with 283 and 119 deaths, respectively).

And based on recovered patients, Ghana had the highest recovered rate (97.6% with 43,693 recovered). The next 9 countries were Cameroon (94.1% with 18,448), Cote d□Ivoire (93.8% with 17,323), South Africa (88.1% with 561,204), Nigeria (78% with 42,922), Egypt (77.4% with 77,208), Morocco (76.9% with 53,929), Algeria (70.5% with 32,481), Kenya (60.4% with 21,158), and Ethiopia had the lowest recovered rate (36.2% with 20,776). These highest rates may indicate that the countries have well protected and have taken strong measures on the virus, but still needs more efforts to control the pandemic in the next long and short times. But, the two East African countries Kenya and Ethiopia have the lowest recovered rates as compared with the others. And, currently the two countries have the highest active cases rates with 37.9% and 62.3%, respectively and the virus is increasing alarmingly as compared with other countries. And, they need strong interventions and treatment centers/hospitals, especially Ethiopia will be open more health facility centers since she has the lowest hospital beds per thousand (0.3) as Who reports showed. And as Who African region report, Ethiopia requires more tests and test reagents in order to intensify implementation of the new strategy for COVID-19 response because of the increase in cases in the country [3–8].

This study will be a guide in what direction the existing healthcare services should increase the countries’ capacities in the coming days in the face of the expected number of COVID-19 new cases. The epidemic’s rapid spreading reveals the necessity of doing what should be done immediately and by taking the right steps.

This study was designed to give communities and also the government a sense of how fast this pandemic is progressing and to inform them of necessary precautions. For this purpose, the number of COVID-19 new cases data of the top 10 infected African countries (South Africa, Egypt, Morocco, Ethiopia, Nigeria, Algeria, Ghana, Kenya, Cameroon, and Côte d’Ivoire) has been modeled and forecasted using the curve estimation regression models and time series models from February 14 and September 6, 2020 in this study. The start date of the virus varies in countries, and thus the models are evaluated and installed separately for each country. The predictions show how the course of the epidemic new cases will be in the following days, taking into account increase rates of the current cases. And, the main motivation of the study was to model the COVID-19 new cases which increase cubically and exponentially in a short time according to the health and social policies of the countries. As a result of modeling, different times dependent policies can be developed based on the estimated and forecasted curves of COVID-19 new cases will be increased, decreased or constantan for the next days. These fitted models will be guiding both in health and social terms. And, these specified models to be published early; they will be a very serious guide for policymakers in the countries.

## 2. Methods

### 2.1. Data Set

The data in this study sets involve the COVID-19 new cases in the top 10 infected African countries from February 14 to September 6, 2020. And, it was downloaded from Our World in Data COVID-19 database and from Worldometer information [3]. [https://github.com/owid/covid-19-data, & https://www.worldometers.info/coronavirus].

### 2.2. Curve Estimations Regression Models

In this study, the fitted modeled and curve estimations models were made for the countries COVID-19 new cases in the given period. The analyses were conducted by SPSS and STATA software. In the study, all the missing values were not included in the analysis.

All the curve estimation regression models for Ln (COVID-19 new cases) are given below:

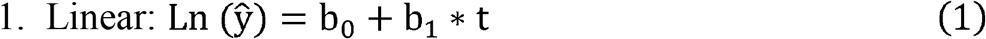

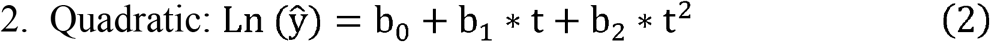

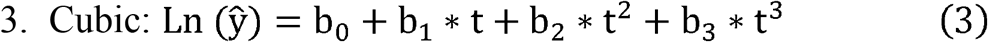

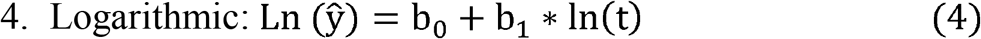

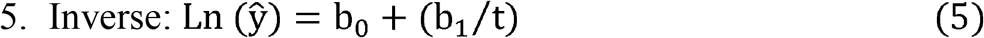

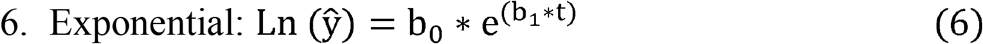

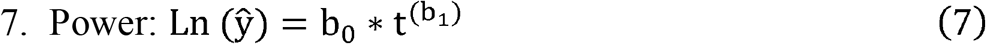

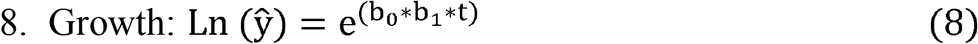

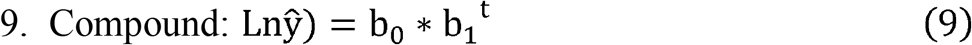

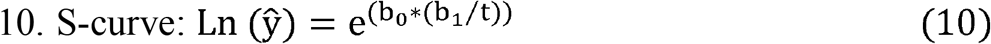

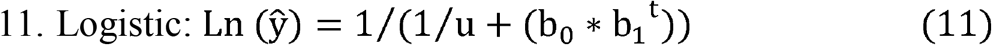

Where: u is the upper boundary whose value is a positive number and greater than the largest dependent variable value.

The fitting effectiveness the models and curves are based on R-squared and standard error of the estimate (SEE). But, the AIC and residual sum-of-squares or SEE are more useful to pick best model then plot the curve to visualize the fit than R^2^ in the nonlinear regression model And, the smallest standard error of the estimate indicates that the data points are closer to the fitted values [11].

### 2.3. Time Series Models

The time series modeling is to carefully collect and rigorously study the past observations of a time series to develop an appropriate model which describes the inherent structure of the series. The series enables to improve a proper model and to make prospective estimations by using statistical methods. However, stationary series are required for estimating the values which they will take prospectively by using the past values for any series. Since non-stationary series contain up-and-down values exhibiting variance at high level, margin of error in the possible estimates is quite high. Stationary may be defined as a probabilistic process whose average and variance do not vary over time and covariance between two periods is based on distance between two periods, not period for which this covariance is calculated methods are used for searching the stationary. Those that are most common among these methods are ACF (Autoregressive Correlation Function) and PACF (Partial Autoregressive Correlation Function) graphics [12–14].

#### 2.3.1. ARIMA Models

Statisticians George Box and Gwilym Jenkins developed a practical approach to build ARIMA model, which best fit to a given time series and also satisfy the parsimony principle. In ARIMA models, a non-stationary time series is made stationary by applying finite differencing of the data points. The ARIMA models include the most suitable but limited parameter among the various model options, depending on the nature of the considering data. ARIMA (p,d,q) models are obtained by taking the difference of series from d degree and adding to ARMA (p, q) model for the stabilizing process. In the ARIMA (p,d,q) models, p is the degree of the Autoregressive (AR) model, q is the degree of the moving average (MA) model and d stands how many differences are required to make the series stationary. ARIMA model becomes AR (p), MA (q) or ARMA (p, q) if the time series is stationary.

The ARMA (p,q) model is shown as follows:

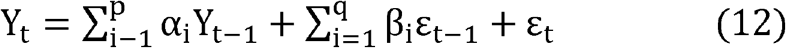

First difference of the non-stationary Y_t_ times-series is observed by (13) below

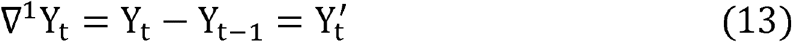

If 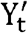 series is not still stationary, difference taking process is repeated for the d times until being stationary. The general form for the difference taking process is given as follows:

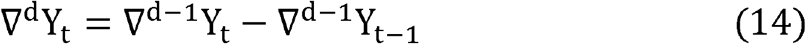

The expression of ARIMA (p, d, q) model can be defined as follows:

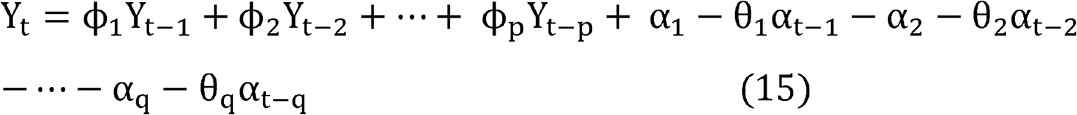

Here: cppare the parameter values for autoregressive operator, αq are the error term coefficients, θqare the parameter values for moving average operator, and Yt is the time series of the original series difference at the degree d [12–15].

#### 2.3.2. Exponential Smoothing Methods

There are four types of non-seasonal exponential smoothing models. Those are Simple, Holt’s linear trend, Brown’s linear trend, and Damped trend models [12, 16–19].

1. Simple model: It is used for forecasting a time series when there is no trend or seasonal pattern, but the mean (or level) of the time series *Y*_*t*_ is slowly changing over time. The simple exponential smoothing model is given by the model equation:

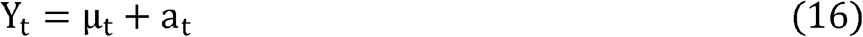

The smoothing equation is

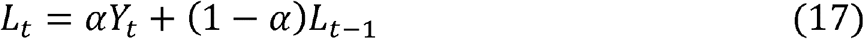

The h-step-ahead prediction equation is

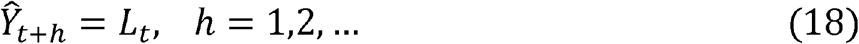

That is the forecast Y h-steps ahead by using the last available estimated (smoothed) level state, L_t_. The ARIMA model equivalent to the simple exponential smoothing model is the

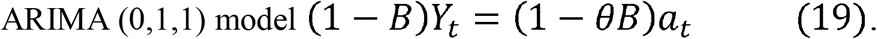

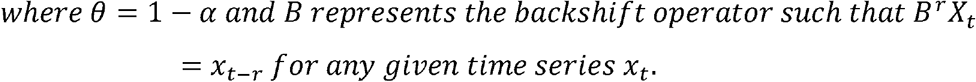
2. Holt’s linear trend model: This model is appropriate for a series with a linear trend and no seasonality. Its relevant smoothing parameters are level and trend, and, in this model, they are not constrained by each other’s values. Holt’s exponential smoothing is most similar to an ARIMA with zero degree of autoregressive, two degrees of differencing, and two degrees of moving average. In this method, estimates are made using the equations below.

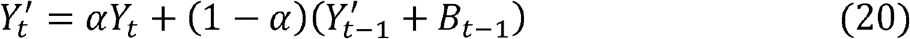

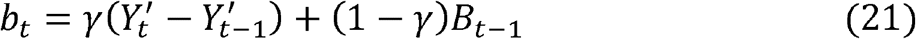

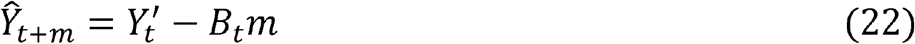

Where α and γ are the smoothing constants in the range of [0, 1].
3. Brown’s linear trend model: It is a special case of Holt linear trend model. In this model, the parameters are assumed that the level and trend are equal. In this method, estimates are made using the equations below.

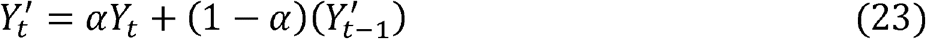

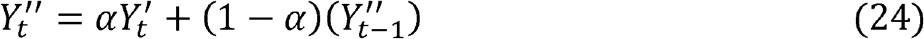

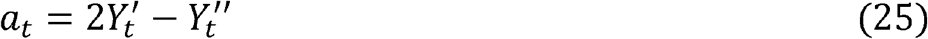

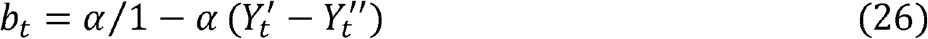

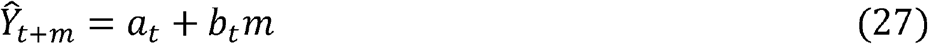
4. Damped trend model: It is a non-seasonal exponential smoothing model which has performed well in numerous empirical studies, and it was well established as an accurate forecasting method. The new stated damped trend model is written as follow:

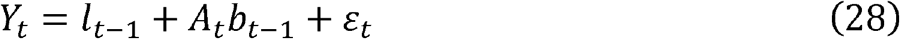

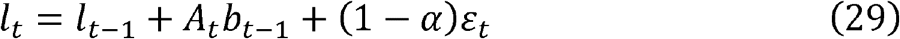

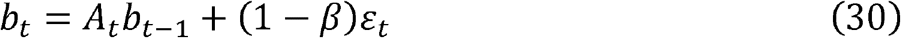

Where Y_t_ is the observed series, l_t_ is its level and b_t_ is the gradient of its linear trend. This model has a single source of error, εt. This model reduced form is a random coefficient ARIMA (1,1,2).

## 3. Results and Discussions

### 3.1. Prevalence of COVID-19 Cumulative Cases

Figure 1 showed that the monthly prevalence of COVID-19 cumulative cases was declined in South Africa, Cote d□Ivoire, Egypt, Ghana, Cameron, Nigeria, and Algeria by 31%, 26%, 22%, 20%, 14%, 12%, and 4% from July to August, respectively. But, it was raised in Ethiopia, Morocco, and Kenya by 41%, 38%, and 1% from July to August, respectively. Specifically in Ethiopia and Morocco, the countries 60% and 54% of the cumulative cases have recorded in August month only, respectively. This indicates that more laboratory tests were conducted in the two countries. Whereas, the two highly infected countries (South Africa and Egypt) have recorded only 22% and 5% of the countries cases in August month, respectively.

**Figure 1:**
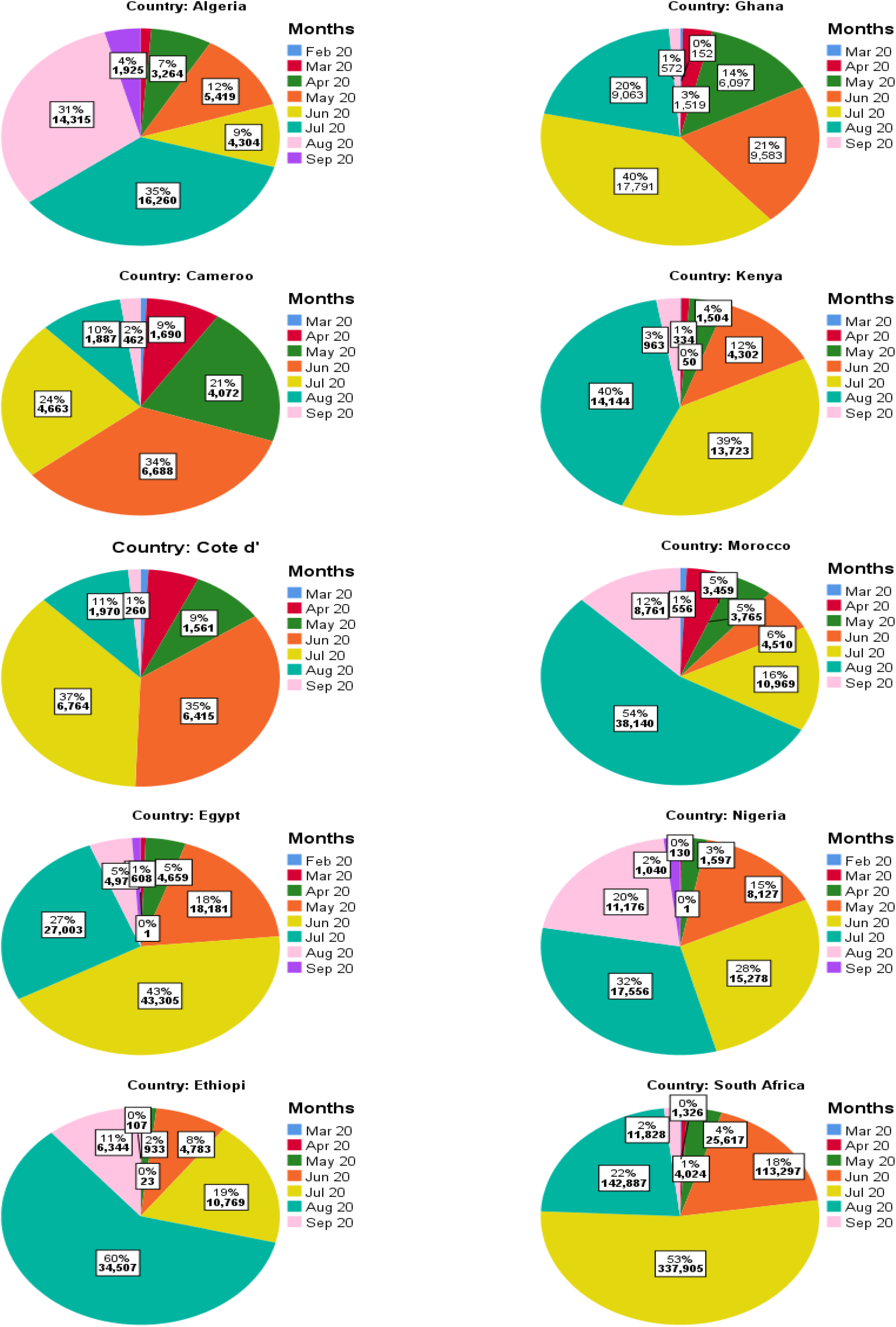
The Pie charts for COVID-19 cumulative new cases by months of the countries.

**Figure 2:**
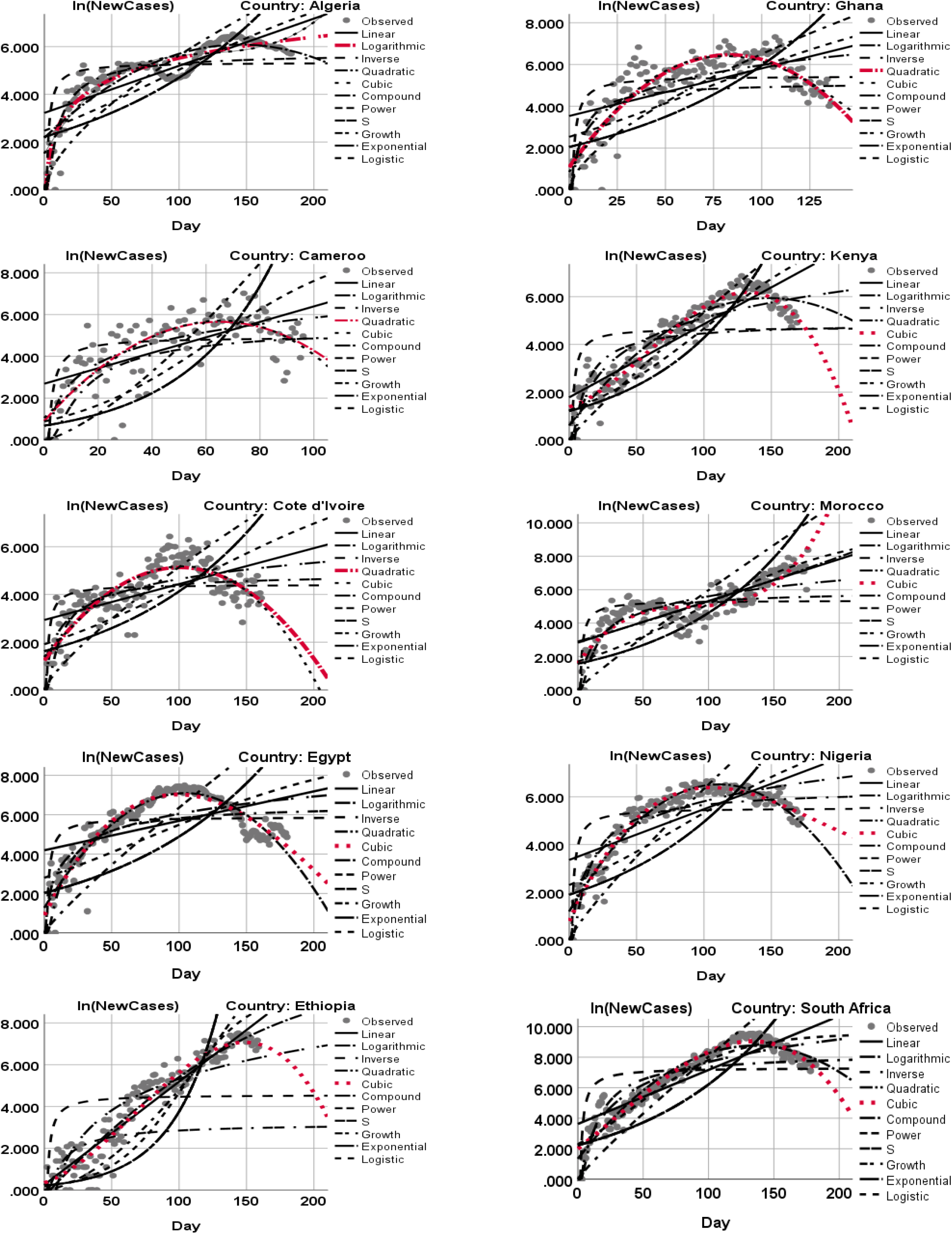
Curve estimates for the countries COVID-19 new cases from Feb 14 to Sep 6, 2020.

**Figure 3:**
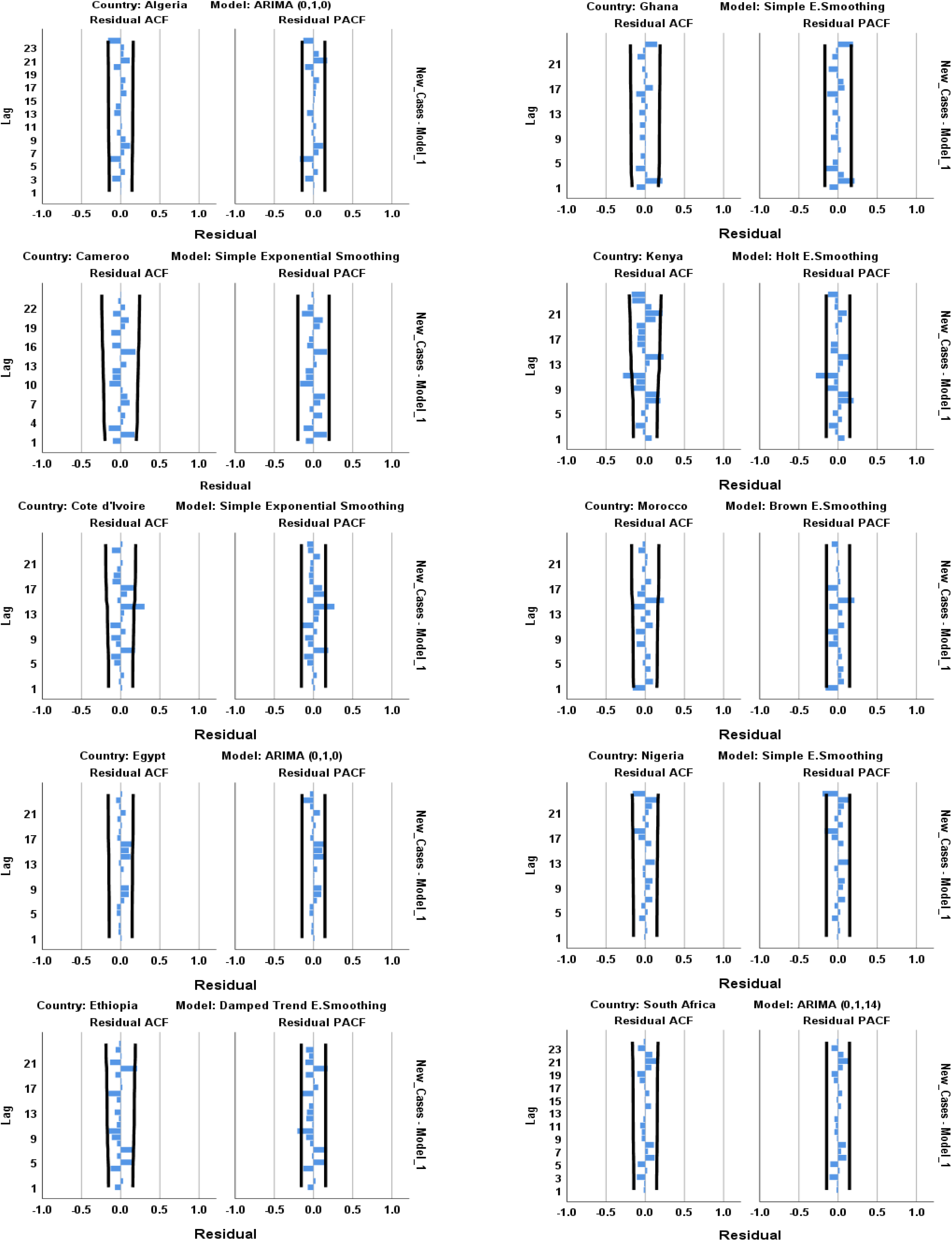
The graphs of ACF and PACF residuals.

**Figure 4:**
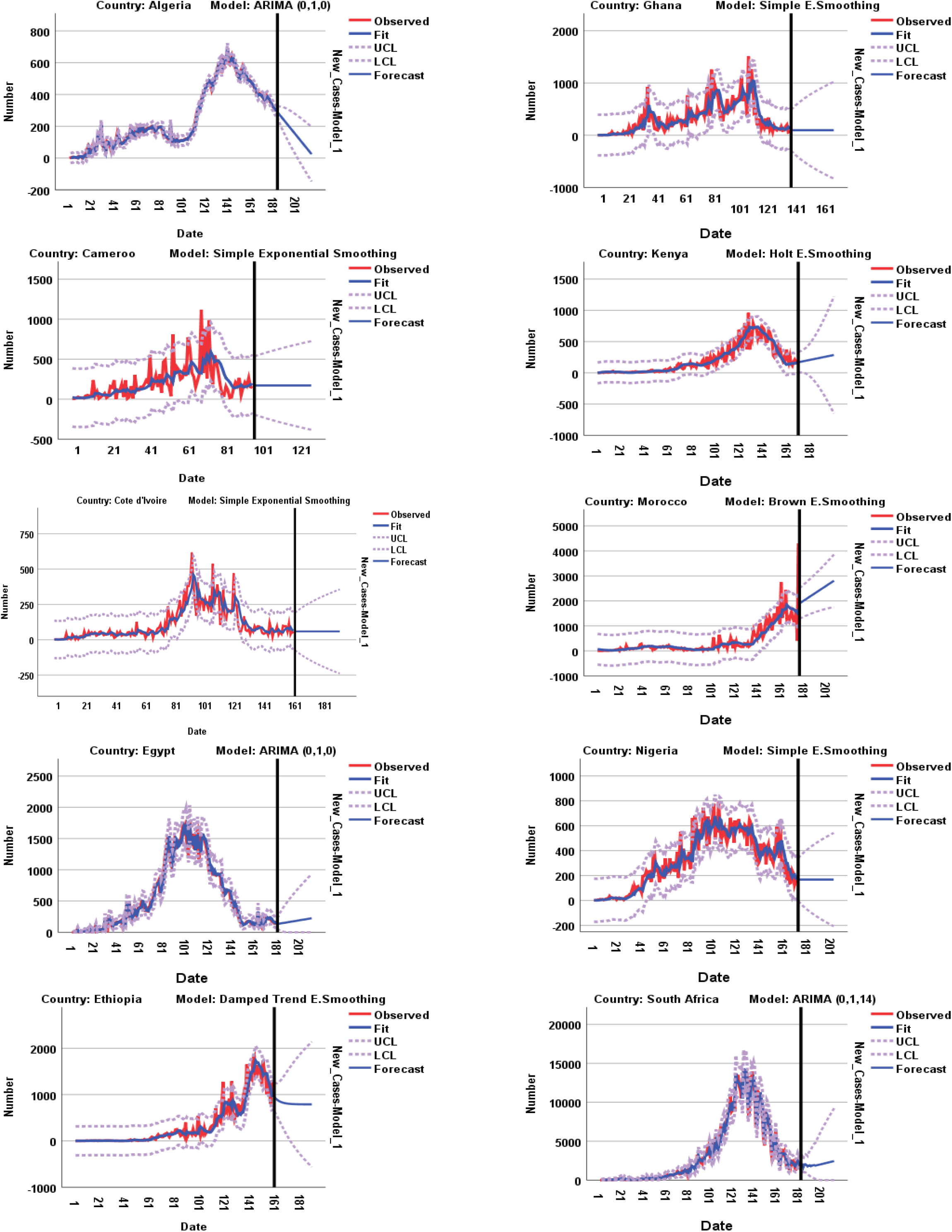
The fitting models and forecasts graphs of COVID-19 new cases.

### 3.2. Regression Fitted Models Summary

The model summary, the fitted models, and the curve estimation graphs were given in table 2 and figure 1 to determine which model fits the data better. Among the regression models, the model with the highest R^2^ and the smallest standard error of the estimate (SEE) values was the best fitted model for the countries COVID-19 new cases data. Thus, the cubic regression model was the best for Egypt, Ethiopia, Kenya, Morocco, Nigeria and South Africa new cases data. And, the quadratic regression model was the best for Cameroon, Cote d□Ivoire and Ghana new cases data. And, Algeria data was followed the logarithmic regression model. The transformed COVID-19 new cases to Ln (new cases) data were more fitted the models and the curves.

**Table 2:**
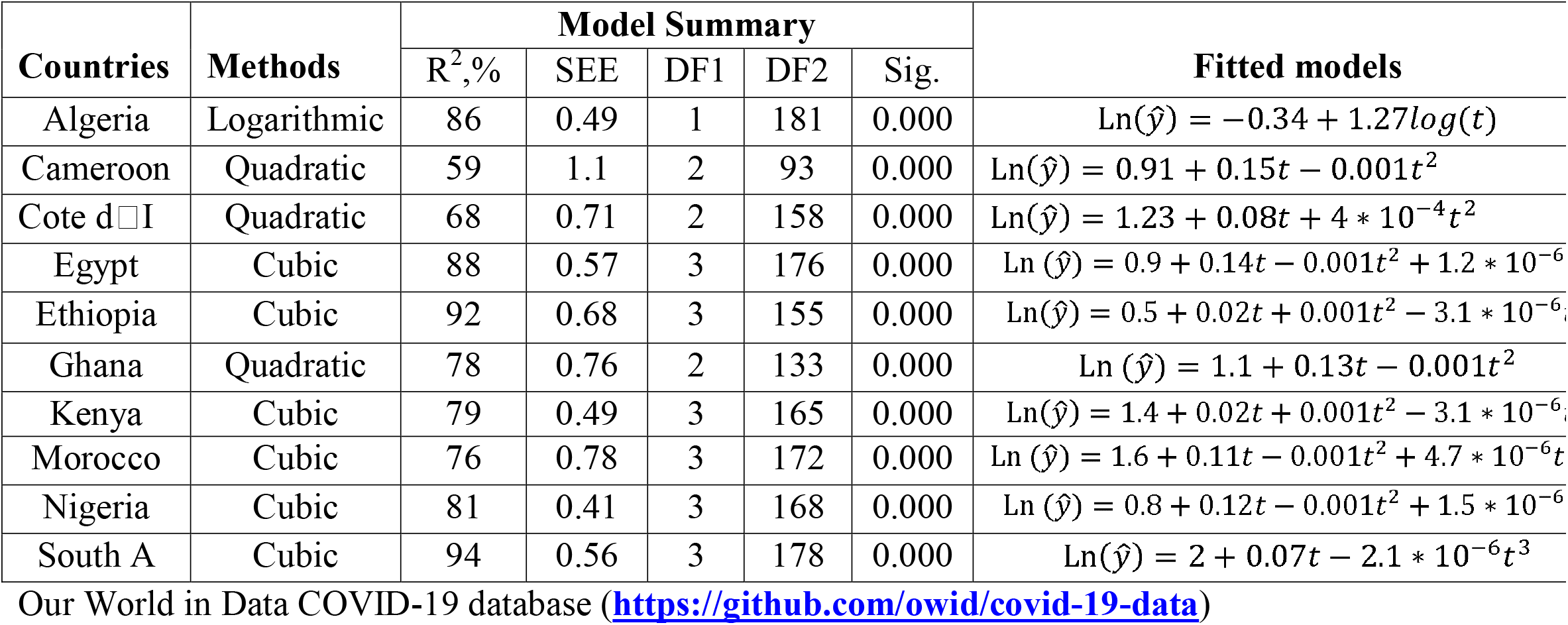
Summary of the fitted regression models of COVID-19 new cases for the countries.

### 3.3. Time Series Fitted Models Summary

The stationary of the residuals is examined and the ACF and PACF graphics of the series for countries are given in Figure 3. When the graphs are examined, there are only a few values that exceed the confidence limit, thus the series can be evaluated as stationary. Table 3 and figure 3 show the goodness of fit criteria values of the exponential smoothing and ARIMA models. And, the appropriate model selection is based on normalized BIC with the smallest value. The Algeria, Egypt, and South Africa COVID-19 new cases data have fitted the ARIMA (0,1,0), ARIMA (0,1,0), and ARIMA (0,1,14) models, respectively. The Cameroon, Cote d’Ivoire, Ghana, and Nigeria data have fitted the same model type (Simple exponential smoothing model). And, the Ethiopia, Kenya, and Morocco data have followed the Damped trend, Holt, and Brown exponential smoothing models, respectively. All of these fitted models have relatively the smallest normalized BIC, root mean square error and mean absolute percentage error values with the highest stationary R-squared and R-squared values. And the Cote d’Ivoire, Ethiopia, Kenya, and Morocco data were statistically significant at 5% level of significance.

**Table 3:** The goodness of fit criteria values for the ARIMA and exponential smoothing models for the countries COVID-19 new cases from February 14 to September 6, 2020.

| Country | Model Type | Model Fit statistics |  |  |  |  | Ljung-Box Q(18) |  | # of Outliers |
| --- | --- | --- | --- | --- | --- | --- | --- | --- | --- |
|  |  | Stationary R<sup>2</sup> | R<sup>2</sup> | RMSE | MAPE | Normalized BIC | DF | Sig. |  |
| Algeria | ARIMA(0,1,0) | 0.638 | 0.994 | 15.655 | 14.669 | 5.788 | 18 | 0.576 | 9 |
| Cameroon | Simple | 0.438 | 0.292 | 183.529 | 191.19 | 10.472 | 17 | 0.193 | 0 |
| Cote d'Ivoire | Simple | 0.272 | 0.679 | 66.815 | 56.825 | 8.435 | 17 | 0.000 | 0 |
| Egypt | ARIMA(0,1,0) | 0.564 | 0.977 | 82.233 | 30.387 | 9.081 | 18 | 0.601 | 8 |
| Ethiopia | Damped Trend | 0.387 | 0.899 | 158.068 | 53.397 | 10.222 | 15 | 0.033 | 0 |
| Ghana | Simple | 0.333 | 0.578 | 196.806 | 53.78 | 10.601 | 17 | 0.412 | 0 |
| Kenya | Holt | 0.787 | 0.861 | 84.317 | 57.394 | 8.93 | 16 | 0.000 | 0 |
| Morocco | Brown | 0.725 | 0.721 | 310.653 | 184.81 | 11.507 | 17 | 0.005 | 0 |
| Nigeria | Simple | 0.304 | 0.84 | 87.487 | 28.188 | 8.973 | 17 | 0.553 | 0 |
| South A | ARIMA(0,1,14) | 0.518 | 0.974 | 680.084 | 91.046 | 13.303 | 14 | 0.464 | 4 |

### 3.4. Forecasts and Trends of COVID-19 New Cases

The forecasted values and trends of COVID-19 new cases data are presented in table 4 and figure 4 for the countries for the next one month from Sep 7 to Oct 6, 2020. The trends will be declined for Algeria and Ethiopia. The trends will be constantan for Cameroon, Cote d’Ivoire, Ghana and Nigeria. But, the trends will be raised slightly for Egypt, Kenya, Morocco, and South Africa.

**Table 4:**
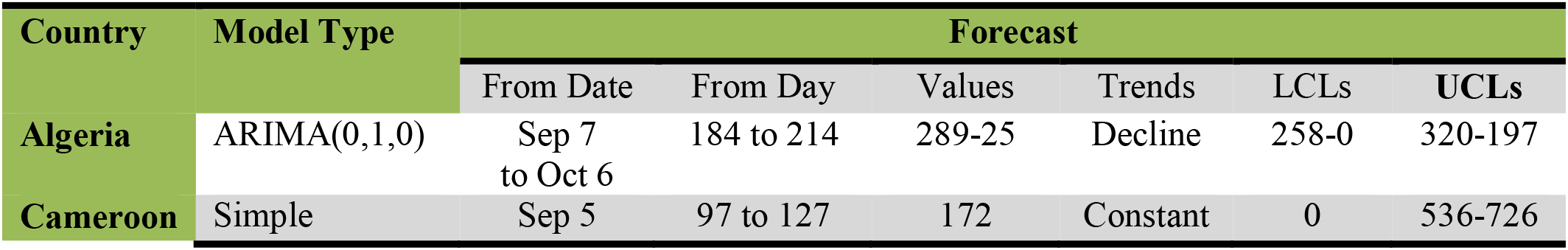
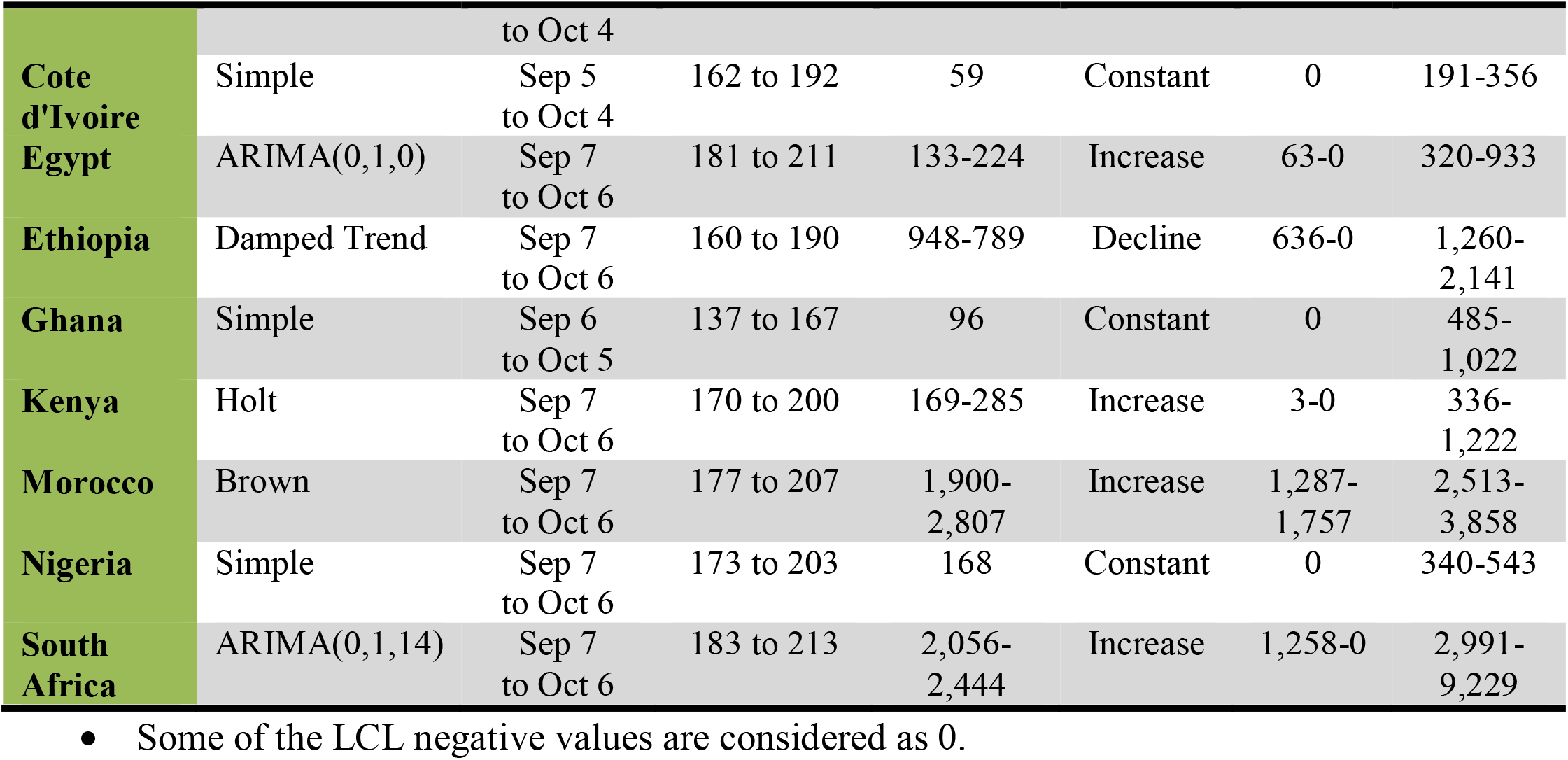
Forecast and trends of COVID-19 new cases for the next one month for the countries.

## 3.5. Discussions

In this study, from July to August, the prevalence of COVID-19 cumulative cases was declined in South Africa, Cote dIvoire, Egypt, Ghana, Cameron, Nigeria, and Algeria by 31%, 26%, 22%, 20%, 14%, 12%, and 4%, respectively. But, it was highly raised in Ethiopia and Morocco by 41%, and 38% in this period, respectively. In Kenya, it was raised only by 1%. In this study the findings from curve estimation regression models for Ln transformation of COVID-19 new cases data showed the cubic regression models were relatively the best fit for Egypt (R^2^=88% with SEE=0.57), Ethiopia (R^2^=92% with SEE=0.68), Kenya (R^2^=79% with SEE=0.49), Morocco (R^2^=76% with SEE=0.78), Nigeria (R^2^=81% with SEE=0.41) and South Africa (R^2^=94% with SEE=0.56). And, the quadratic regression models were the best fit for Cameroon (R^2^=59% with SEE=1.1), Cote dIvoire (R^2^=68% with SEE=71) and Ghana (R^2^=78% with SEE=0.76). And, Algeria data was followed the logarithmic regression model (R^2^=86% with SEE=0.49). All the models were statistically significant at 1% level of significance.

A similar study in the selected G8 European countries (Germany, United Kingdom, France, Italy, Russian, Canada, Japan, and Turkey) for the number of COVID 19 epidemic cases data were fitted the cubic regression models with the curve estimations. And, all these cubic fitted models were statistically significant at 1% level of significance [20].

And, another study using predictive models (polynomial regression models) were considered based on the WHO daily rampant data. The result of the study had shown that the spatial and temporal pattern of this novel virus was varying, spreading and covering the entire world within a brief time. In the study, the fitting effect of cubic model (R^2^=99.6%) was a best outperforming compared to the other six families of exponentials [21].

The time series modeling of this study found that the Algeria, Egypt, and South Africa COVID-19 new cases data have fitted the ARIMA (0,1,0), ARIMA (0,1,0), and ARIMA (0,1,14) models, respectively. The Cameroon, Cote d’Ivoire, Ghana, and Nigeria data have fitted the same model type (Simple exponential smoothing model). And, the Ethiopia, Kenya, and Morocco data have followed the Damped trend, Holt, and Brown exponential smoothing models, respectively. But, only the Cote d’Ivoire, Ethiopia, Kenya, and Morocco models were statistically significant at 5% level of significance. In this study, the forecasted values and trends of COVID-19 new cases also presented for each country for the next one month from Sep 7 to Oct 6, 2020. The trends will be declined for Algeria and Ethiopia from 289 to 25 with 95% CL of 289-197 and from 948 to 789 with 95% CL of 636-2,141, respectively. The trends will be constantan for Cameroon, Cote d’Ivoire, Ghana and Nigeria through 172 with 95% CL of 0-726, 59 with 95% CL of 0-356), 96 with 95% CL of 0-1,022 and 168 with 95% CL of 0-543, respectively. Whereas, the trends will be raised slightly for Egypt from 133 to 224 with 95% CL of 63-933, slightly for Kenya from 169 to 285 with 95% CL of 3-1,222, significantly for Morocco from 1,900 to 2,807 with 95% CL of 1,287-3,858, and significantly for South Africa from 2,056 to 2,444 with 95% CLs of 1,258-9,229.

The study in the selected G8 European countries (Germany, United Kingdom, France, Italy, Russian, Canada, Japan, and Turkey) for the number of COVID 19 epidemic cases data were modeled and forecasted that Japan (Holt Model), Germany (ARIMA (1,4,0) and France (ARIMA (0,1,3) were provided statistically significant. The UK (Holt Model), Canada (Holt Model), Italy (Holt Model) and Turkey (ARIMA (1,4,0) were not statistically significant [20].

A study in Africa also showed that the spatial pattern of cumulative COVID-19 cases in Morocco was the leading contributor to the burden of COVID-19 in Northern African in June 30, 2020. Morocco had forecasted 4,459,877 (CI 95% 689,547 to 28,845,720) cumulative cases of COVID-19 and this was almost double the estimated number for Algeria, a country with the next highest burden, 2,804,674 (CI 95% 438,629 to 17,935, 144) by the end of June 2020. In Southern Africa, South Africa and Swaziland are the leading contributors to the pandemic. By the end of June 2020, the countries were expected to have 2,581366 (CI 95% 404,835 to 16,459, 651) and 254,403 (CI 95% 45,730 to 1,415,274) cumulative cases, respectively. In the Western Africa sub-region, cumulative cases of infection were dominated by Cote d’Ivoire and Ghana, despite Nigeria having a larger population than both countries combined. And the numbers of new COVID-19 infections were expected to increase from 2,453,700 (CI 95% 418,527 to 14,374,452) cases in April, to 5778830 (CI 95% 967402 to 34,802,668) cases in May, to 8,044,927 (CI 95% 1,326,846 to 49,176,051) cases by the end of July. This represents a 135% increase from April to May and a 39% increase from May to June, which suggests a possible slowing down of the pandemic with time [5].

And another study using African COVID-19 cases found that the estimated the exponential growth rate was 0.22 per day (95% CI: 0.20–0.24), and the basic reproduction number (R_0_) was 2.37 (95% CI: 2.22–2.51) based on the assumption that the exponential growth starting from 1 March 2020. With an R_0_ at 2.37, we quantified the instantaneous transmissibility of the outbreak by the time-varying effective reproductive number to show the potential of COVID-19 to spread across African region [22].

And, other study in Africa region also presented that the epidemic could be basically controlled in late April with strict control of scenario one, manifested by the circumstance in the South Africa and Senegal. Under moderate control of scenario two, the number of infected people will in-crease by 1.43–1.55 times of that in scenario one, the date of the epidemic being controlled would be delayed by about 10 days, and Algeria, Nigeria, and Kenya are in accordance with this situation. In the third scenario of weak control, the epidemic would be controlled by late may, the total number of infected cases would double that in scenario two, and Egypt was in line with this prediction [23].

## 4. Conclusion and Recommendation

### 4.1. Conclusion

From July to August, the prevalence of COVID-19 cumulative cases was declined in South Africa, Cote dIvoire, Egypt, Ghana, Cameron, Nigeria, and Algeria by 31%, 26%, 22%, 20%, 14%, 12%, and 4%, respectively. But, it was highly raised in Ethiopia and Morocco by 41%, and 38% in this period, respectively. In Kenya, it was raised only by 1%.

In this study, the cubic regression models for the ln(COVID-19 new cases) data were relatively the best fit for Egypt, Ethiopia, Kenya, Morocco, Nigeria and South Africa. And, the quadratic regression models for the data were the best fit for Cameroon, Cote dIvoire and Ghana. The Algeria data was followed the logarithmic regression model.

In the time series analysis, the Algeria, Egypt, and South Africa COVID-19 new cases data have fitted the ARIMA (0,1,0), ARIMA (0,1,0), and ARIMA (0,1,14) models, respectively. The Cameroon, Cote d’Ivoire, Ghana, and Nigeria data have fitted the simple exponential smoothing models. The Ethiopia, Kenya, and Morocco data have followed the Damped trend, Holt, and Brown exponential smoothing models, respectively. In the analysis, the trends of COVID-19 new cases will be declined for Algeria and Ethiopia, and the trends will be constantan for Cameroon, Cote d’Ivoire, Ghana and Nigeria. But, it will be raised slightly for Egypt and Kenya, and significantly for Morocco and South Africa from September 7 to October 6, 2020.

The measures taken by countries such as the individual attitudes of the societies towards the specified measures and the number of virus tests to be performed are factors that may affect the number of cases. Since this study was conducted with the current measures, the forecasts obtained may differ from the number of cases that occur in the future.

Thus, the study findings should be useful in preparedness planning against further spread of the COVID-19 epidemic in Africa.

### 4.2. Recommendation

The researcher recommended that as many countries continue to relax restrictions on movement and mass gatherings, and more are opening up their airspaces to international travelers with easing of quarantine measures for returning residents and visitors, and the countries’ different public and private sectors (like Schools, Universities, Stadiums, and others) are reopening. While these are necessary actions, the appropriate public health and social measures must be instituted on the ground. So, these measures include again, but are not limited to, early detection of suspect cases and tracing contacts to confirmed cases, case management, risk communication and keeping up with infection prevention and control guidelines.

In future studies, more data and healthier evaluations can be made as a matter of course. However, since this study provides information about the levels that the number of cases can reach if the course of the current situation cannot be intervened, it can guide countries to take the necessary measures again.

## Data Availability

In this study, the available data are attained based on the daily reported cases of the WHO, GitHub (owid-COVID-19 data) and Worldometer.

## Funding

The author received no financial support for the research, authorship, and publication of this article.

### Acknowledgements

Thanks to WHO, GitHub (owid-COVID-19 data) and Worldometer for COVID-19 cumulative and new cases data reports, and also Ambo University for internet access.

## Author’s Contributions

The author’s contribution is reading all articles in this area and analyzing the data released by GitHub and Worldometer, and then make into more meaningful. Therefore, this data and all the ideas are done by the author alone.

## Ethics approval and consent to participate

In this study, the research considers data released by GitHub (owid-COVID-19 data).

## Consent for publication

This paper is purely done based on the data released by GitHub (owid-COVID-19 data) and all information related to others articles are already cited as references.

## Trial Registration

The trial registration of this study considers data related to GitHub (owid-COVID-19 data) and Worldometer that were being COVID-19 cumulative cases and new cases. Therefore, all relevant information could be obtained from these sites. The results were not published therefore it is retrospectively registered.

